# Trans-Ancestry GWAS of Hot Flashes Reveals Potent Treatment Target and Overlap with Psychiatric Disorders

**DOI:** 10.1101/2024.09.12.24313572

**Authors:** Kathryn E. Werwath, Rebecca B. Lawn, Madeleine T. Salem, Tayden Li, Brittany L. Mitchell, Hanyang Shen, Scott D. Gordon, Benson Kung, Ciera Stafford, Mytilee Vemuri, Andrew Ratanatharathorn, Joeri Meijsen, Aladdin H. Shadyab, Charles Kooperberg, Karestan C. Koenen, Carolyn J. Crandall, Nicholas G. Martin, Laramie E. Duncan

**Affiliations:** Department of Psychiatry and Behavioral Sciences, Stanford University, Stanford, CA, USA; Department of Epidemiology, Harvard T.H. Chan School of Public Health, Harvard University, MA, USA; Queensland Institute of Medical Research, Brisbane, Queensland, Australia; Institute for Biological Psychiatry, Copenhagen, Denmark; Herbert Wertheim School of Public Health and Human Longevity Science, University of California San Diego, La Jolla, CA, USA; Division of Public Health Sciences, Fred Hutchinson Cancer Center, Seattle, WA, USA; Department of Social and Behavioral Sciences, Harvard T.H. Chan School of Public Health, Harvard University, MA, USA; Psychiatric and Neurodevelopmental Genetics Unit, Department of Psychiatry, Massachusetts General Hospital, Boston, MA, USA; Division of General Internal Medicine and Health Services Research, Dept. of Medicine, David Geffen School of Medicine at University of California Los Angeles, Los Angeles, CA, USA

## Abstract

**Background:** Most women experience hot flashes (hot flushes) during the menopause transition. Menopausal hot flashes typically persist for years. For a sizeable minority of women, hot flashes are severe and substantially impairing. It is worthwhile to further investigate the genetic underpinnings of hot flashes.

**Method:** We conducted the largest trans-ancestry genome-wide association study (GWAS) of hot flashes available to date (N=149,560). We used self-assessment of hot flashes in the Nurses’ Health Study, Nurses’ Health Study II, Women’s Health Initiative, and Queensland Institute of Medical Research samples (total n=42,489). In one sample (UK Biobank, n=107,071) direct assessment of hot flashes was not available, so menopausal hormone therapy was used as a proxy variable. We estimated the heritability of hot flashes and genetic correlations with psychiatric phenotypes using linkage disequilibrium score regression (LDSR).

**Results:** In component analyses and our trans-ancestry meta-analysis, the top locus was on chromosome 4 in the neurokinin 3 receptor gene (*TACR3*, position 104,556,732, trans-ancestry *p*=7.2×10^−41^). A second novel locus was identified (*LINC02428, p*=3.5×10^−8^). Gene results implicated *TACR3, GRID1, NUDT4*, and *PHF21B*. Using the hot flash GWAS meta-analysis (n=42,489; i.e., no proxy variable), SNP heritability was estimated: *h*^*2*^_liab_=.08 (*h*^*2*^_SNP_=.04, *se*=.02). Genetic correlations were statistically significant between hot flashes and posttraumatic stress disorder (PTSD, *rg*=0.25, *p*=0.01), schizophrenia (*rg*=0.17, *p*=0.02), and depression (*rg*=0.21, *p*=0.01).

**Discussion:** These genomic findings are consistent with independent, robust basic science research which led to a novel treatment for hot flashes, namely, neurokinin 3 receptor antagonists. This new class of hot flash drugs blocks the receptor (neurokinin 3 receptor) coded for by the top locus for hot flashes (*TACR3*). This GWAS of hot flashes provides an uncommonly clear example of how GWAS findings can point to potent treatment targets for complex brain phenotypes. We also found that the proxy variable (menopausal hormone therapy) pointed to the same target (*TACR3*), and that exclusively intronic and intergenic variants signaled this target.

## Introduction

The experience of a hot flashes includes the sudden onset of heat, typically in the upper body, which may be accompanied by sweating, flushing, and chills^1,2^. Hot flashes, also called hot flushes, are commonly experienced by approximately 70% of women after menopause, but over half of women also experience hot flashes in the years leading up to menopause (i.e., during the late menstrual transition) ^1,3,4^. Menopause is defined as the point at which a woman has not experienced a menstrual period for 12 consecutive months, and the average age of menopause is 51. Though commonly referred to separately, it has been noted that night sweats are likely hot flashes experienced during sleep^2^. Collectively, hot flashes and night sweats are known as vasomotor symptoms. Throughout this manuscript we will use the term hot flashes for simplicity of phrasing and given that the term “vasomotor symptoms” is less well known.

One of the most misunderstood aspects of menopause is the severity and duration of its symptoms, which can persist for many years for a large fraction of women. Accordingly, many women leave the workforce because of menopausal symptoms including hot flashes^5–7^. The economic impact of menopausal symptoms is substantial, with an estimated annual cost of $1.8 billion attributed solely to missed workdays^5^. Historically, menopausal symptoms have been downplayed, ignored, and inadequately treated. Recognizing the importance of addressing these issues, a 2024 executive order allocated $12 billion for Women’s Health Research and Innovation, with a focus on advancing menopause research as a critical component. Given that menopause is a core life stage for women, it is critical to further elucidate the genetic underpinnings of symptoms^5^.

In the first genome-wide association study (GWAS) conducted for hot flashes, Crandall et al. (2017)^8^ found a significant locus in the *TACR3* gene in a trans-ancestry meta-analysis of 17,695 post-menopausal women from the Women’s Health Initiative (WHI). The *TACR3* locus was replicated in a sample from the Study of Women Across the Nation^9^ and women from the UK Biobank with linked primary health care records^10^. Because many samples fail to gather data about hot flashes, some studies have used proxy outcome variables for hot flashes. In previous work using the UK Biobank, we performed a GWAS of estrogen medication usage and identified *TACR3* as the only significant locus^11^.

Given somewhat limited research about the genetics of hot flashes to date, it is particularly noteworthy that prior results are consistent with the known biology of hot flashes. Specifically, the *TACR3* gene encodes the NK3R receptor that binds neurokinin B (NKB, also known as neurokinin 3), and NKB signaling is a causal component in a neural circuit that is sufficient for generating of hot flashes^12^. Evidence supporting the critical role of NKB signaling in hot flashes comes from multiple complementary sources: human post-mortem tissue studies^13,14^, animal studies^12,15^, and mechanistic studies in humans^16^. This body of work led to the development of a novel medication treatment for hot flashes that blocks the NK3R receptor, which was approved by the FDA in 2024 (fezolinetant)^17–21^.

Here, we report the largest GWAS of hot flashes to date, incorporating five cohorts and 149,560 women in a trans-ancestry meta-analysis. In addition to Crandall et al.’s prior GWAS in the WHI, we conducted GWAS in the Nurses’ Health Study (NHS), the Nurses’ Health Study II (NHSII), and in samples from the Queensland Institute of Medical Research (QIMR). We also conducted a GWAS of menopausal hormone therapy (MHT), formerly known as hormone replacement therapy (HRT), use in the UK Biobank as a proxy for hot flashes; this builds upon our prior work using a less specific medication variable as a proxy for hot flashes.^11^ Here we sought to 1) identify novel loci and genes for hot flashes, 2) estimate SNP heritability for hot flashes, and 3) quantify genetic correlations between hot flashes and psychiatric phenotypes.

## Methods

This GWAS meta-analysis includes new GWAS analyses that we conducted (in NHS, NHSII, QIMR, and UK Biobank) and summary statistics from the published GWAS by Crandall et al (for WHI)^8^ Major methodological elements are described below and additional details are provided in the **Supplementary Materials**.

### Samples

Data from 149,560 post-menopausal women were included in this analysis. To be included, women needed to be post-menopausal (per self-report, as is the standard for defining menopause). Women also needed to have answered questions about the experience of having hot flashes and/or night sweats affirmatively or negatively (i.e. women with ambiguous responses were excluded). In the UK Biobank, affirmative or negative responses for the use of MHT were required. Women also needed genome-wide genotype data passing quality control procedures (see below) to be included (see **Supplementary Text** for further details). **Figure 1** presents an overview of sample sizes and ancestries for each cohort; **Table 1** presents sample characteristics. Ethical approval and participant consent were obtained according to each cohort’s institutional protocols.

**Table 1.** Participant and cohort characteristics.

| Study name | N | Ancestry* | Age | Phenotype assessment | Case N (%) | Location |
| --- | --- | --- | --- | --- | --- | --- |
| NHS + NHSII | 24,249 | European | 52-88 | ever experienced hot flashes and/or night sweats | 18,543 (76%) | United States |
| QIMR | 545 | European | 35-82 |  | 462 (85%) | Australia |
| WHI | 17,695 | European<br>Hispanic<br>African | 50-79 |  | 5,157 (63%)<br>5,184 (77%)<br>1,889 (68%) | United States |
| UK Biobank | 107,071 | European<br>South Asian<br>African<br>East Asian | 40-71 | ever used MHT | 44,754 (43%)<br>237 (17%)<br>409 (24%)<br>146 (23%) | United Kingdom |

**Figure 1.**
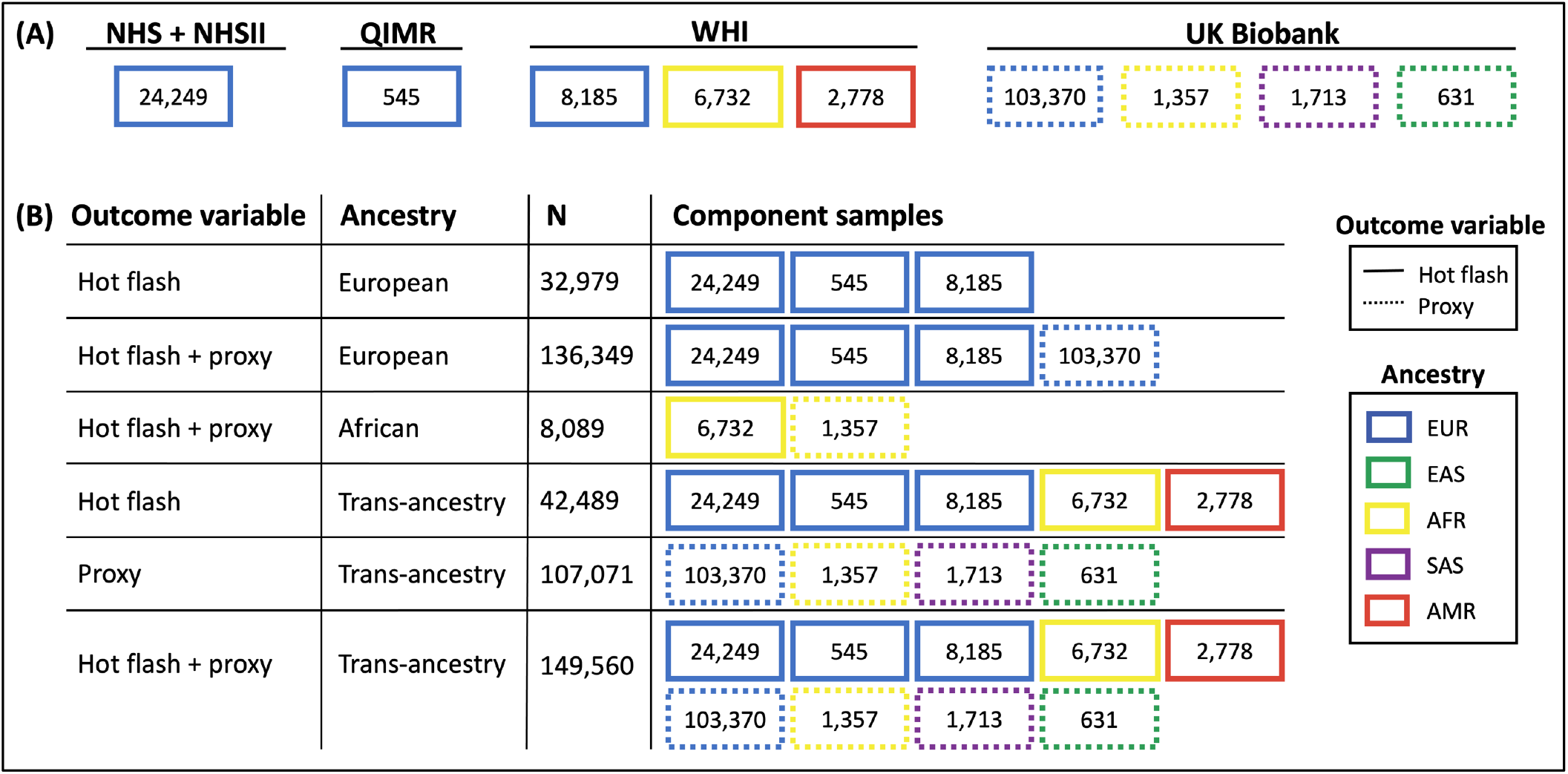
Schematic of component GWAS samples and GWAS meta-analyses. (A) GWAS samples by cohort with sample sizes. Solid borders denote studies that asked women about hot flashes as contrasted with dashed borders denoting the UK Biobank samples that did not ask women about hot flashes, but for whom women reported use (or not) of menopausal hormone therapy (MHT). Border colors represent major distinctions regarding ancestry using 1000 Genomes Project groups as reference^22^. (B) Depiction of each meta-analysis conducted by component samples. NHS=Nurses’ Health Study, NHSII=Nurses’ Health Study II, QIMR=Queensland Institute of Medical Research, WHI=Women’s Health Initiative, EUR=European ancestry, EAS=East Asian ancestry, AFR=African ancestry, SAS=South Asian ancestry, AMR=Hispanic/Latino ancestry.

### Phenotypic Measures

Across all samples, the outcome phenotype was a binary yes/no variable denoting any experience of a hot flash or night sweat, and in the UK Biobank a binary yes/no variable denoting use of MHT. In NHS, experiences of hot flashes were queried in a 2008 questionnaire which asked women to report whether “at the beginning of menopause, did you have hot flashes or night sweats? (if you took estrogen, consider the time period before starting the treatment)”. The same question regarding hot flashes was asked in NHSII participants in 2009, 2013, and 2017. In the UK Biobank baseline assessment (2006-2010), women were asked whether they had ever used MHT (with no further specifications, e.g., about oral vs. transdermal, estrogen with or without a progestogen); women who responded “do not know” or “prefer not to say” were set to missing and therefore not included in the sample definition. In QIMR, women were drawn from two sources and queried either from 1989-1992 or in 1996. The questions, respectively, were: “Please tell us if you had any of the following symptoms of menopause that women sometimes experience: hot or sudden flushes or sweats”^23^ and “number and intensity of hot flushes and accompanying sweats, often at night?”^24^ Questions were answered on a 1-4 frequency scale with 1 denoting “not at all”. Responses were re-coded as a binary variable (yes/no, for experiencing hot flashes) to better match coding of data from the other cohorts.

### Genotyping, imputation, quality control

Genotyping was performed using blood samples collected from 2006-2010 in UK Biobank, 1989-1990 in NHS, 1996-1999 in NHSII, and from 2009 onwards in QIMR^8,23–27^. In brief, data for our samples were genotyped using two arrays in the UK Biobank (UK BiLEVE, UK Biobank axiom array), five in NHS (Illumina HumanHap Array, Illumina OncoArray, Illumina HumanCore Exome Chip, Illumina OmniExpress, Affymetrix 6.0), and three in NHSII (Illumina HumanHap Array, Illumina OncoArray, Illumina HumanCore Exome Chip). QIMR used HapMap-based (370K, 610K, 660K) or 1000-Genomes-based (Core+Exome, PsychArray) Illumina SNP arrays. Genetic data were imputed to the Haplotype Reference Consortium (HRC) reference panels^28^ and UK Biobank data was also imputed using the UK10K panel. Standard GWAS quality control procedures were conducted (see **Supplementary Material** for details).

### Statistical analyses

#### Software summary

We conducted genome-wide association studies using logistic regression in PLINK 2.0^29^ and in SAIGE 0.45 (QIMR only). Meta-analyses of GWAS results were conducted using METAL (version released 2011-03-25)^30^. Power calculations were conducted using the genpwr package^31^ in R (v4.0.5)^32^. Mendelian Randomization was conducted using MRBase^33^. R (v4.0.5)^32^ was used for other analyses and to create figures. Locus Zoom was used for the SNP Manhattan plot^34^. Biorender was used to create neuron depictions.

#### GWAS and meta-analyses

In NHS and NHSII cohorts, GWAS was conducted separately for each array and by batch for data from the Illumina HumanCore Exome Chip in NHSII^25^. In the UK Biobank, GWAS was conducted for each ancestry group (see **Supplementary Figure S1** for ancestry assignment). All GWAS were adjusted for principal components according to norms in each dataset (WHI=10, NHS=10, QIMR=4, and UK Biobank=20). GWAS in the UK Biobank were additionally adjusted for the Townsend deprivation index and, in European ancestry analyses, the difference in years between each participant’s age at menopause and 2002 (to account for potential effects of WHI findings about MHT, released in 2002, on women’s likelihood of using MHT). Note that we found no evidence of differences in genetic effect sizes related of timing of menopause before or after 2002, when the paradigm-changing WHI findings were released^35^, and thus we opted not to include this extra covariate in the other UK Biobank GWASs, which were much smaller. GWAS in QIMR included covariates for imputation run (see **Supplementary Material** for details). The chromosome and position of SNPs is reported here according to genome build GRCh37/hg19.

#### Meta-analyses

We ran multiple rounds of meta-analyses across outcome phenotypes and ancestry groups (see **Figure 1** and **Supplementary Material**). Our main analysis incorporated the largest number of women (n=149,560), thus including all ancestries available (trans-ancestry) and both phenotypic outcomes (hot flashes and the MHT proxy). For all meta-analyses, we used METAL (version released 2011-03-25).^30^ A fixed-effects meta-analysis with inverse variance weighting was conducted. Note that no additional loci were identified when we conducted a variable effects meta-analysis.

#### Post-hoc power calculations for component samples

We sought to determine whether we had the power to detect the known *TACR3* locus in component samples. Power calculations were conducted using the genpwr package^31^ in R (v4.0.5)^32^ with the corresponding sample size, case rate, and minor allele frequency for the component samples. The estimated effect size for the *TACR3* locus was *OR*=1.63 for hot flashes and *OR*=1.18 for the MHT proxy. Alpha was set at *p*<5×10^−8^.

#### Gene-level analyses

Gene p-values were obtained with MAGMA (v1.10)^36^ and plots were generated using R (v.4.0.5)^32^.

#### SNP heritability and genetic correlations

We used linkage disequilibrium score regression (LDSC)^37^ to estimate SNP heritability and to examine genetic correlations with psychiatric phenotypes. We used the meta-analysis of hot flashes in European ancestry samples (n=32,979; see **Figure 1**) for these analyses because LDSC requires non-admixed ancestry for samples, and because this was the best powered analysis (see power calculations). For the genetic correlations, we used publicly available GWAS data for posttraumatic stress disorder (PTSD)^38^, depression^39^, bipolar disorder^40^, schizophrenia^41^, and Alzheimer’s disease^42^. We further included a sensitivity analysis in which *h*^*2*^_*SNP*_ and genetic correlations were estimated after removing the top locus (chromosome 4:104,424,934-104,801,645).

#### Mendelian Randomization

We used Mendelian Randomization (MR) to gain insight into possible causal relationships between hot flashes and psychiatric phenotypes. In MR, SNPs that are strongly associated with a trait of interest serve as instruments to estimate the causal effect of that trait on other phenotypes. For a SNP to be considered a valid instrument in MR, it must at least satisfy three main conditions: i) the SNP is associated with the exposure trait of interest; ii) it affects the outcome solely through that exposure; and iii) it is not linked to any third factor that influences both the exposure and outcome, thereby avoiding horizontal pleiotropy. Instrumental variables for hot flashes were chosen from those present across traits examined by clumping genome-wide suggestive SNPs using PLINK (*p*<5×10^−6^, 10,000 Kb and *r2*<0.001). The causal relationship between trait and hot flashes was tested using two-sample MR with the MRBase package^33^, employing inverse-variance weighted (IVW) MR, which combines a Wald-type estimator for each instrument in a multiplicative random effects model. To ensure consistency in the results, sensitivity analyses were performed using four additional MR methods: MR Egger, Weighted Median, and Weighted Mode. In brief, MR-Egger is similar to the IVW method but adjusts for directional horizontal pleiotropy of the SNPs used as instruments; the weighted median accounts for up to half of the instrument SNPs having a pleiotropic effect; and the weighted mode is the mode of the Wald-type estimates used in the IVW, weighted by standard error-based weights for each SNP. As a sensitivity analysis, all MR analysis was repeated using the hot flash GWAS excluding the *TACR3* locus.

### Data availability

Cohort data may be made available upon reviewed requests from WHI, NHS, and UK Biobank, and further details relevant to this project are provided in the **Supplementary Text**. Genome-wide summary data from the present study can be downloaded from XXX.

### Extended analyses

During the course of work on this project, Ruth et al. (2023)^10^ published a GWAS of hot flashes using UK Biobank data. This analysis included 92,028 women, with hot flashes reported in linked primary care records as a phenotype. They included only European ancestry individuals. We used the summary statistics from Ruth et al. to compare and extend our results. Specifically, we compared effect sizes of our top locus with Ruth et al., and we used the Ruth et al. dataset in place of our UK Biobank results to conduct an alternative meta-analysis and gene-level analyses.

## Results

### GWAS and GWAS meta-analyses

**Figure 2A** presents the Manhattan plot for our hot flash only (i.e., no proxy) trans-ancestry meta-analysis (n=42,489), in which two loci reached genome-wide significance. **Figure 2B** presents the Manhattan plot for our hot flash + MHT proxy trans-ancestry meta-analysis (N=149,560), which has one additional significant locus. Overall, we found the strongest association for rs74827081, which lies in the *TACR3* gene on chromosome 4 (*p*=7.2×10^−41^, *OR=*1.27, 95% *CI*=1.23-1.32, position 104,556,732). This SNP has the highest posterior probability of causality for this locus (0.517, per LocusZoom)^34^. All significant (*p*<5×10^−8^) SNPs in this locus are in introns of the *TACR3* gene or upstream of *TACR3*. Nearby GWAS Catalog hits include age at menarche and testosterone concentrations^43^. Note that the effect size for rs74827081 (*OR*=1.27) from the hot flash + MHT proxy meta-analysis is in between the effect size for hot flashes (*OR*=1.63, *95% CI*=1.52-1.75) and the MHT proxy (*OR*=1.18, *95% CI*=1.13-1.23), consistent with expectation of an attenuated effect size for the proxy variable (i.e. 1.63 to 1.18). See **Figure 3** for a forest plot of effect sizes. For top SNP rs74827081(G/C), the C allele increases the risk of hot flashes and likelihood of MHT use.

**Figure 2.**
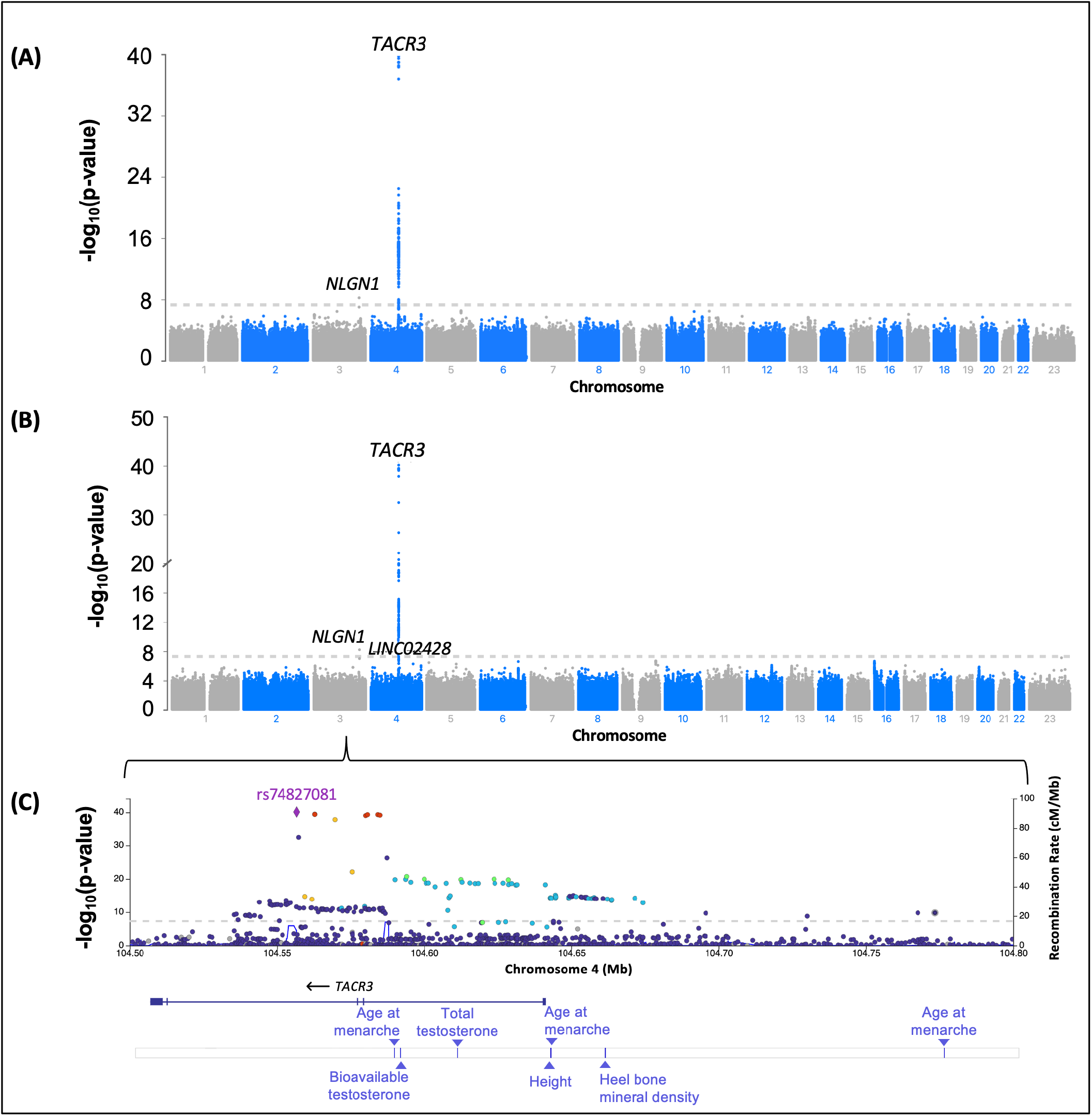
SNP level results for trans-ancestry meta-analyses. (A) Manhattan plot showing statistical significance for each SNP (-log_10_(*p*)) for the hot flash only (i.e., no proxy) trans-ancestry meta-analysis (n=42,489). (B) Manhattan plot for the hot flash + MHT proxy trans-ancestry meta-analysis (N=149,560). (C) Region plot with color denoting linkage disequilibrium between the top SNP (rs74827081) and nearby SNPs. The GWAS catalog hits bar shows previously reported associations in this region. LD=linkage disequilibrium, Mb=megabase, cM=centimorgan.

**Figure 3.**
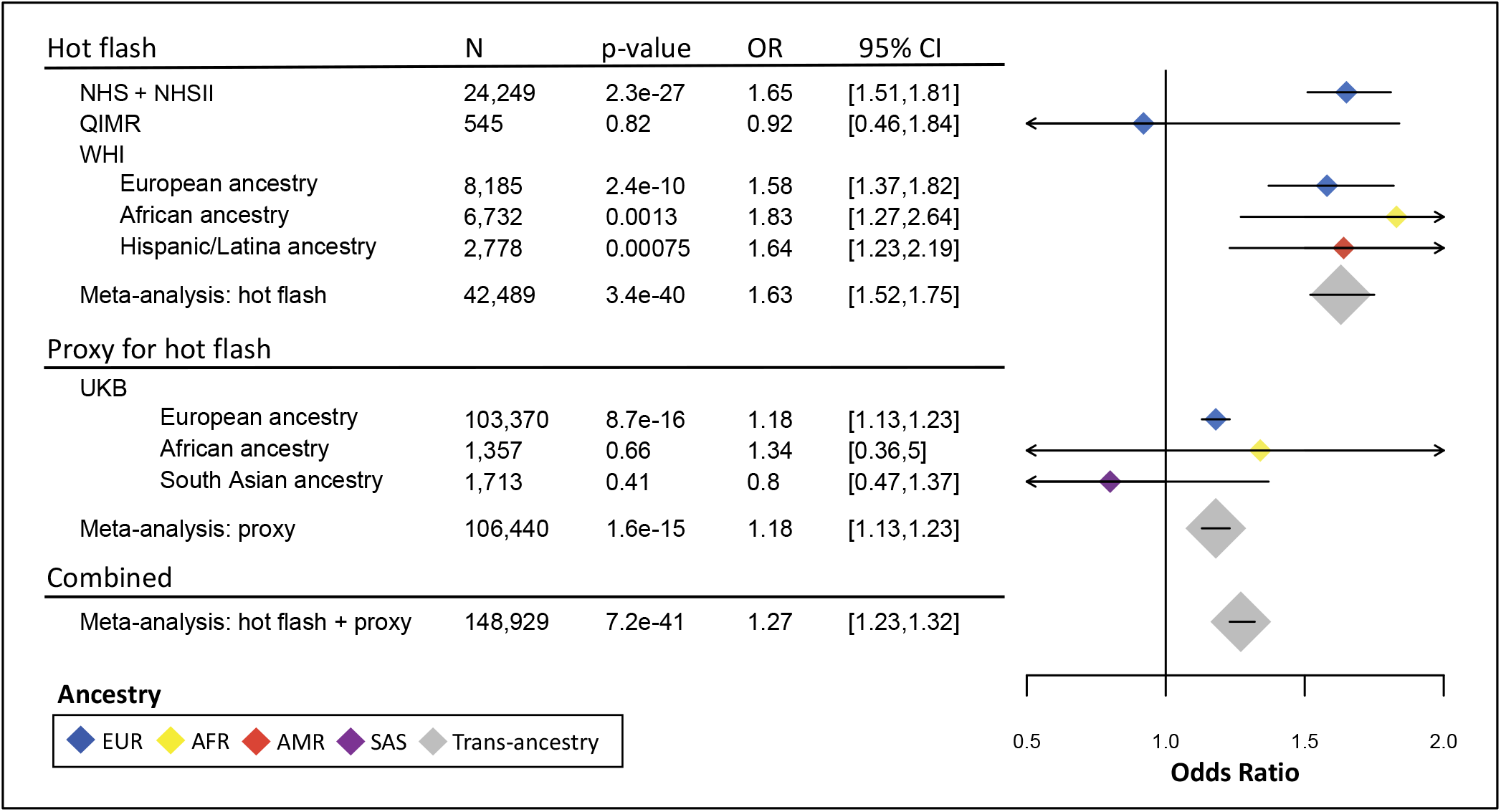
Forest plot of effect sizes for top SNP rs74827081. Odds ratios are plotted as diamonds. Large diamonds denote meta-analytic results as contrasted with small diamonds for component samples. Color denotes ancestry according to 1000 Genomes Project classifications, and grey represents trans-ancestry meta-analyses.^22^ Each odds ratio is plotted along with its 95% confidence interval (as needed, arrows indicate that the interval extends beyond the range of the plot). Samples are grouped by outcome variable. The first grouping includes all hot flash samples. The second grouping includes all samples with the MHT proxy variable. The third is the combined meta-analysis of hot flash and proxy samples. Please note that the final sample size (n=148,929) is smaller than the full sample size (N=149,560) because this locus was not available in the East Asian subsample (n=631).

Regarding other loci (**see Figure 2B**), we found that rs148680409 in the *NLGN1* gene was genome-wide significant (*p*=6.1×10^−9^). However, as the SNP was only measured in the WHI data, we could not confirm this result in any other samples, and this is not considered a high confidence result. The third locus was in the *LINC02428* gene (with top SNP rs13107507, *p*=3.5×10^−8^). This SNP is 0.3 Mb away (located at 4:104244426) from the top SNP but may represent a distinct signal given there are multiple recombination peaks in between the two SNPs (pairwise LD *r*^*2*^=.08). One further SNP on the X chromosome, located at 23:123,080,030, was close to reaching genome-wide significance (*p*=7.6×10^−8^), and note that X chromosome data was only available for the UK Biobank and QIMR datasets. In gene-level analysis, we identified two other genes besides *TACR3* that were significantly associated with hot flashes: the *GRID1* gene on chromosome 10, and the *NUDT4* gene on chromosome 12 (see **Figure 4**) and see **Supplementary Tables S1** and **S2**.

**Figure 4.**
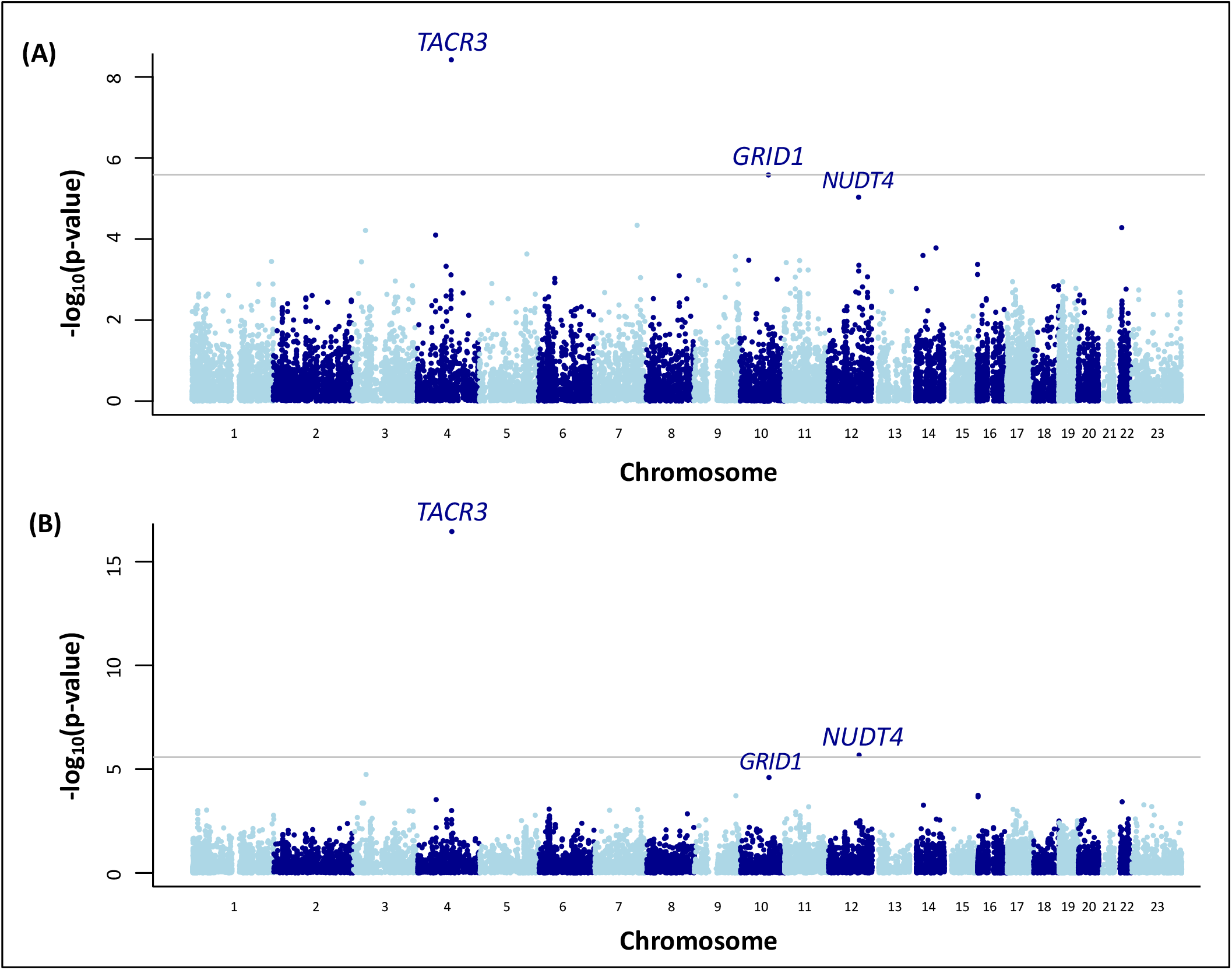
Gene-level results. Gene Manhattan plots for (A) UK Biobank European ancestry MHT proxy sample (n=103,370) and (B) hot flash + MHT proxy European ancestry meta-analysis (n=136,349).

To demonstrate real-world correlates of the effect of *TACR3* genotypes on women’s likelihood of developing hot flashes, we obtained the prevalence of hot flashes by rs74827081 genotype in the NHS and NHSII datasets, to which we had access to individual level data (see **Figure 5**). In this sample, 57% of women with two protective alleles experienced hot flashes (CC), as contrasted with 72% of women with one risk allele (CG), and 80% of women with two risk alleles (GG). Using population allele frequencies from gnomAD^44^, we note that the “high-risk” genotype (GG) is the most common globally; this genotype ranges in frequency from approximately 89% in European ancestry women to approximately 100% in East Asian women.

**Figure 5.**
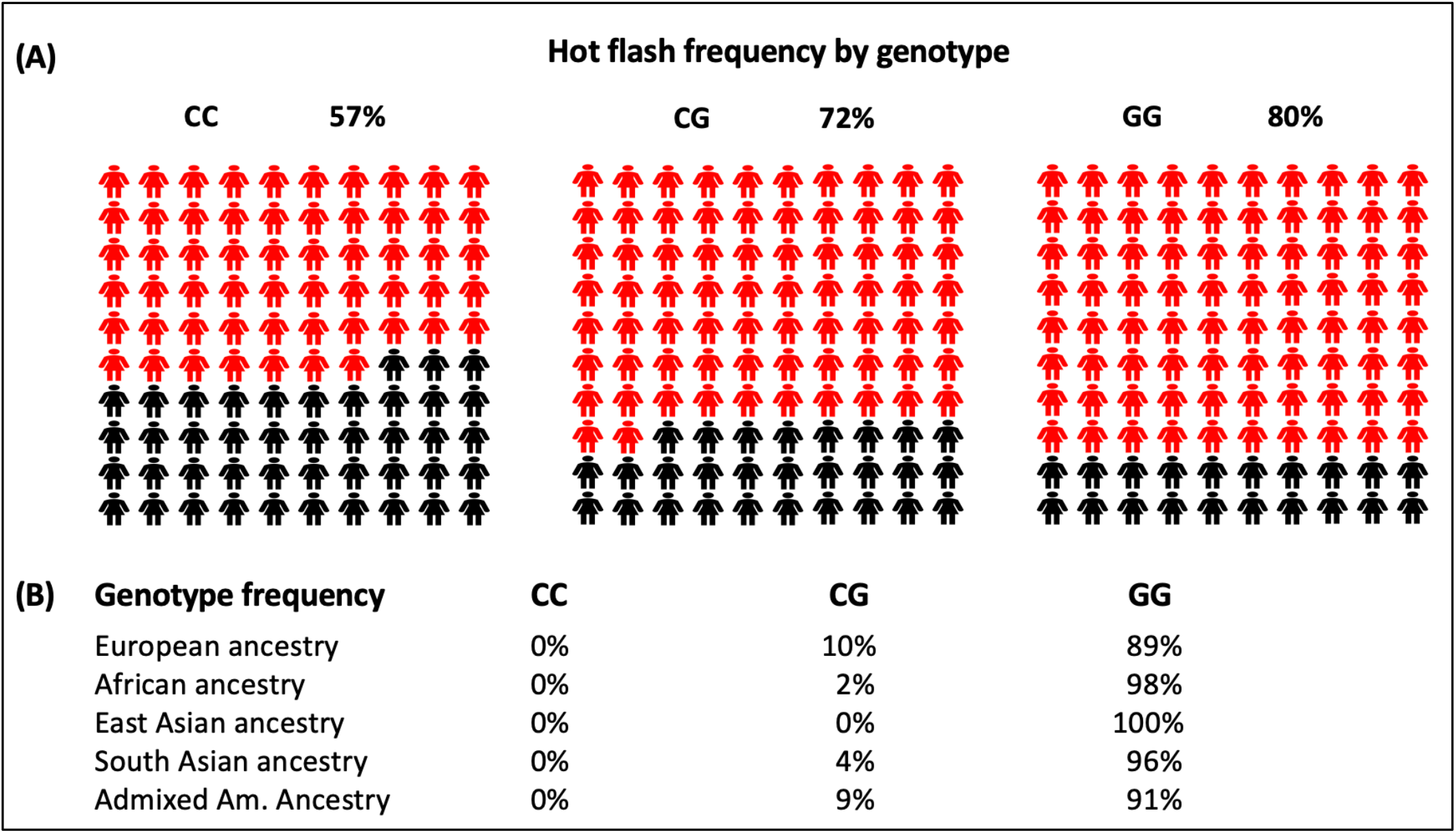
Real world implications of natural variation in the *TACR3* gene. (A) Percentage of women who experienced hot flashes (shaded in red) as a function of rs74827081 genotype in NHS and NHSII. (B) The frequency of each genotype across ancestries as calculated using frequencies from the gnomAD database.

### Post-hoc power calculations

Power calculations for component datasets demonstrate that significant effects were identified as expected according to statistical power. As shown in **Table 2**, at alpha=5×10^−8^, power was very low (less than 5%) for all datasets in which we failed to detect the *TACR3* locus (including all non-European ancestry samples), showing that this is a sufficient explanation for the samples that yielded non-significant results. However, using alpha=.05 for the WHI African ancestry sample and the WHI Hispanic/Latino ancestry sample, power was 69% and 91% respectively, and indeed, we found nominally significant associations for rs74827081 in these samples (*p*=.0012 and *p*=.00075, respectively).

**Table 2.** Statistical power to detect *TACR3* rs74827081 associations in component analyses. At alpha=5×10^−8^, statistical power is a sufficient explanation for the pattern of significant and non-significant results for the *TACR3* locus. For the East Asian sample in the UK Biobank, genotypes were not available, and so the frequency of this variant in gnomAD samples was used (i.e., minor allele frequency=0.0003877).

| Sample | Ancestry | N | Case Rate | MAF | Power (%) | Genome-wide significance for *TACR3* locus |
| --- | --- | --- | --- | --- | --- | --- |
| NHS + NHSII | European | 24,249 | 0.63 | 0.06 | ~100% | yes |
| QIMR | European | 545 | 0.63 | 0.06 | 0.01% | no |
| WHI | European | 8,185 | 0.63 | 0.06 | 91.00% | yes |
| WHI | African | 6,732 | 0.77 | 0.01 | 0.15% | no |
| WHI | Hispanic/Latino | 2,778 | 0.68 | 0.05 | 1.89% | no |
| UK Biobank | European | 103,370 | 0.43 | 0.06 | ~100% | yes |
| UK Biobank | African | 1,357 | 0.17 | 0.01 | 0.00% | no |
| UK Biobank | South Asian | 1,713 | 0.24 | 0.02 | 0.00% | no |
| UK Biobank | East Asian | 631 | 0.23 | 0.00 | 0.00% | no |

### SNP Heritability, genetic correlations, and Mendelian Randomization

Using LDSC, the estimated SNP heritability for hot flashes was *h*^*2*^_SNP.liability_=.08 on the liability scale (*h*^*2*^_SNP.observed_=.04, *se*=.02). Removing the *TACR3* locus, the heritability estimate was very similar *h*^*2*^_SNP.liability_=.08 (*h*^*2*^_SNP.observed_=.04, *se*=.02). Using LDSC, we found significant (p<0.05) genetic correlations between hot flashes and PTSD (*rg*=.25, *se*=.09, *p*=.01), depression (*rg*=.21, *se*=.08, *p*=.01), and schizophrenia (*rg*=.17, *se*=.07, *p*=.02); see **Figure 6**. The genetic correlations changed very little when the *TACR3* locus was removed (solid vs. dashed bars; all SNP in this window removed for the latter: chromosome 4:104,424,934-104,801,645). This suggests that the shared genetic etiology between hot flashes and these other phenotypes is largely independent of the *TACR3* locus and is instead attributable to more diffuse polygenic contributions shared between hot flashes and these other phenotypes. After correcting for multiple testing, none of the Mendelian Randomization results were significant (see **Supplementary Table S3**).

**Figure 6.**
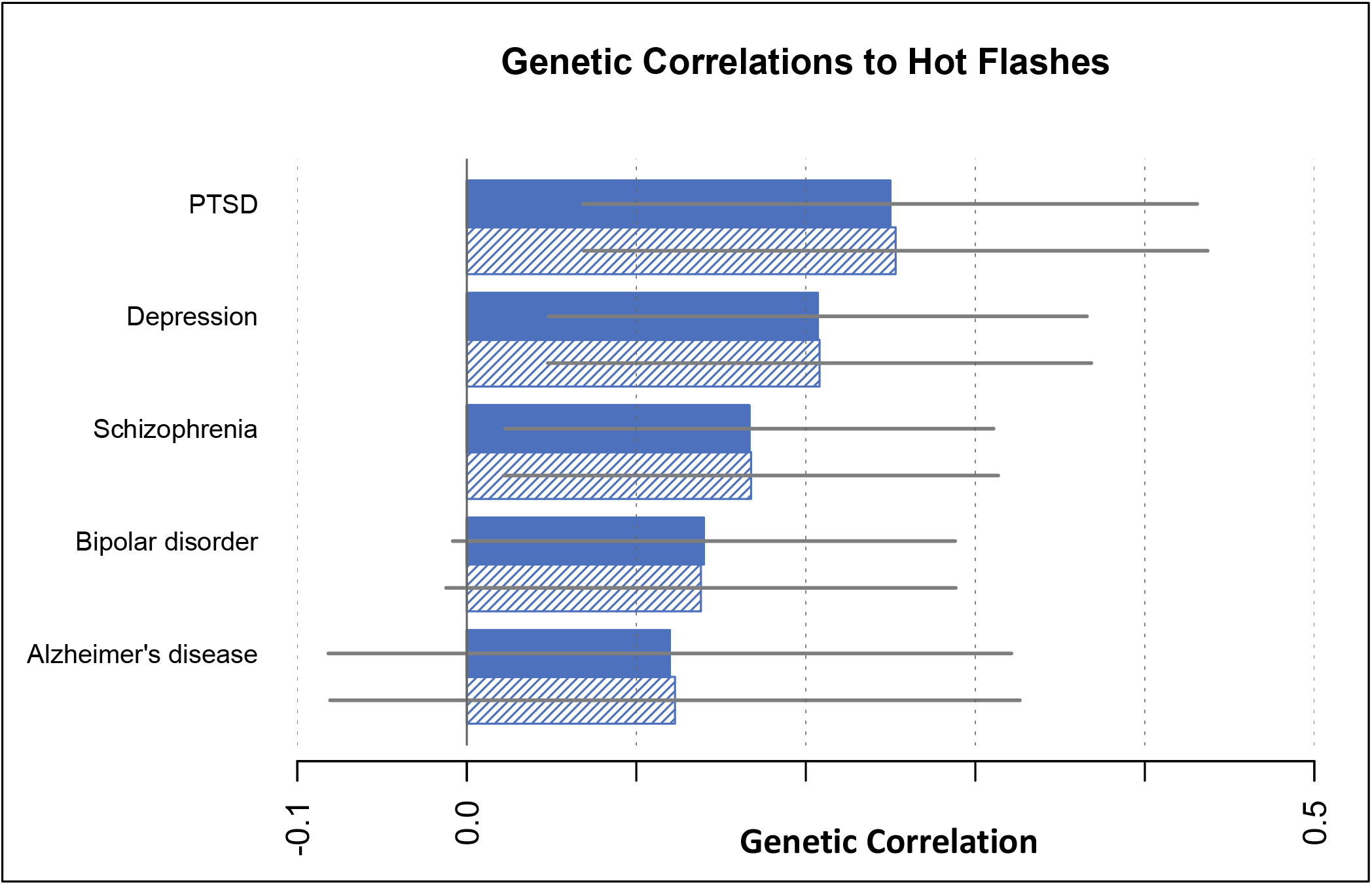
Genetic correlations between hot flashes and psychiatric phenotypes. Genetic correlations were calculated between the hot flash European ancestry meta-analysis (n=32,979) and the listed phenotypes. Each genetic correlation is plotted as a bar along with its 95% confidence interval. Each phenotype has two bars: the solid bar is the genetic correlation with the hot flash meta-analysis including all loci, while the striped bar excludes the top locus (4:104556732). PTSD: posttraumatic stress disorder.

### Extended analyses

Our top SNP (rs74827081) in the Ruth data had an effect size of 1.28 (*p=*2.8×10^−20^, *95% CI=*1.22-1.36), which is between that of our proxy meta-analysis (*OR=*1.18) and our hot flash meta-analysis (*OR=*1.63). This suggests that Ruth’s outcome variable, hot flashes reported in linked primary care records, is a better proxy for hot flashes than self-reported MHT use, but still is not equivalent to self-reported hot flashes. We performed an alternative hot flash + proxy trans-ancestry meta-analysis, with Ruth et al. replacing our UKB European ancestry data (n=138,218). The *TACR3* locus became even more significant (*p=*2.7×10^−52^) compared to the original hot flash + MHT proxy trans-ancestry meta-analysis (*p*=7.2×10^−41^). We also performed an alternative gene-level analysis, with Ruth et al. replacing our UKB data in the European ancestry hot flash + proxy meta-analysis (n=125,007), see **Supplementary Table 4** for full results and **Supplementary Figure S2**. *TACR3* remained significant (*p=*1.8×10^−13^) and a new gene, *PHF21B*, was also significant (*p=*2.1×10^−7^).

## Discussion

In this study, we conducted the largest GWAS of hot flashes to date and identified a highly significant locus in the *TACR3* gene. We also identified three new putative genes for hot flashes (*GRID1, NUDT4*, and *PHF21B*) and several psychiatric phenotypes that have significant genetic correlations with hot flashes. Interestingly, these genetic correlations with multiple psychiatric disorders do not depend on the *TACR3* locus. Rather, it is other polygenic contributions to hot flashes that overlap with genetic risk for psychiatric disorders.

Our main finding in the *TACR3* gene is remarkably consistent with basic science research and with clinical trials data for a new hot flash medication. Specifically, prior research found that the neurokinin B (NKB) receptor in the hypothalamus plays a causal role in hot flash generation. This preclinical work led to the development of an NKB receptor antagonist that is highly effective at treating hot flashes^16–21^. Given that our top locus is in the *TACR3* gene, and that the *TACR3* gene encodes the NKB receptor, we see that this GWAS of hot flashes points directly towards a potent treatment target.

In addition to the *TACR3* locus, we also identified a novel locus in the *LINC02428* gene. Nearby catalog hits for this SNP include apolipoprotein A1 levels, HDL cholesterol levels, and male puberty timing (age at voice breaking)^43^. The *LINC02428* gene codes for long intergenic non-protein coding RNA, which is involved in the liver and various cancers^45,46^. Further research is needed to determine how this locus may contribute to hot flashes. Also of note was an nearly significant SNP on the X chromosome. This locus did not intersect any genes (closest protein coding gene *STAG2*). Given that menopause is a sex-restricted phenomenon, including X chromosome data will be particularly important for future analyses of hot flashes and other menopausal symptoms.

We sought to identify loci for hot flashes in non-Europeans ancestry groups. Our analysis included GWAS results from six non-Europeans ancestry sub-samples, and we conducted the first African ancestry meta-analysis. Unfortunately, power to detect the largest known effect size for hot flashes (in *TACR3*) was very low in these samples (i.e. <2% statistical power). This was not only due to smaller sample sizes, but also to the lower minor allele frequency for the risk allele in non-European ancestry populations. As an example, though the African Ancestry meta-analysis had a sample size of 8,089, which is very close to the WHI European ancestry sample size of 8,185 that was sufficient for the detection of the *TACR3* locus, the minor allele frequency was ∼1% in African ancestry samples as compared to European ancestry samples ∼6%. Continuing to collect multi-ancestry data is a priority for future work in this space.

Gene level analyses identified the *GRID1, NUDT4*, and *PHF21B* genes as significant in European ancestry meta-analyses. The *GRID1* gene codes for a glutamate receptor subunit, and the top tissues for *GRID1* expression include several regions of the brain (including the hypothalamus), as well as the thyroid and uterus^47–49^. The *NUDT4* gene is involved in intracellular trafficking, and the top tissues for *NUDT4* expression include the arteries, heart, esophagus, and ovary^47–49^. Given expression in this set of tissues, it is possible that this gene may play a role in the vasodilation that occurs during a hot flash^47–49^. The top tissues for *PHF21B* include the cervix, pituitary, and vagina^47–49^. The data used for describing tissue-level expression of these genes was obtained from the GTEx Portal on 8/26/24.

We identified several psychiatric phenotypes that have significant genetic correlations with hot flashes. The directions of these genetic correlations are consistent with epidemiological findings. For example, women with depression have been found to be more likely to experience hot flashes^50^, and the genetic correlation findings reported here suggest that shared genetic risk factors may partially explain this above-chance comorbidity. Regarding PTSD, both trauma and PTSD symptoms have been found to be associated with higher risk of hot flashes^51^. Shared genetic influences on hot flashes and psychiatric disorders may help to explain why antidepressant medications are helpful for some women experiencing hot flashes.

Regarding limitations, it is true that MHT is used for symptoms other than hot flashes. It is also only used by a minority of women, whereas most women experience hot flashes. For these reasons, MHT use is an imperfect proxy for hot flashes. Consistent with this, the effect size for the top locus for the MHT proxy was much smaller than the effect size for hot flashes (OR=1.18 vs. 1.63, with non-overlapping 95% confidence intervals). Nevertheless, the proxy variable used here (MHT) yielded the same top locus (*TACR3*). Similarly, we found that the phenotype used by Ruth et al.,^10^ hot flashes reported through linked primary care records, was better than the MHT proxy but was still not equivalent to self-reported hot flashes (OR=1.28). Nevertheless, including Ruth et al. in our alternative trans-ancestry meta-analysis increased the significance of the *TACR3* locus by ten orders of magnitude.

In sum, to our knowledge, this is the largest trans-ancestry GWAS of hot flashes to date. We discovered a novel locus in the *LINC02428* gene. Moreover, our top locus in the *TACR3* gene points directly to an effective treatment target for a complex brain phenotype. Specifically, this locus is in the neurokinin 3 receptor gene, and a neurokinin 3 receptor antagonist (fezolinetant) was recently FDA approved for the treatment of moderate to severe hot flashes^52^. We also identified several new potential genes for hot flashes (*GRID1, NUDT4*, and *PHF21B*), and found significant genetic correlations between hot flashes and psychiatric phenotypes.

## Data Availability

Data availability is dependent upon regulations specific to each of the datasets analyzed here. None are publicly available without approval. Rather, relevant access procedures must be followed to obtain access to datasets including the UK Biobank.

## Acknowledgements

We thank all participants for making this work possible. This work was supported by the Jaswa Innovator Award to LD and by the National Institute of Mental Health (NIMH) to LD (R01 MH123486 & R21 MH125358). We thank the participants and staff of the NHS and NHSII for their valuable contributions and acknowledge the Channing Division of Network Medicine, Department of Medicine, Brigham and Women’s Hospital, and Harvard Medical School for managing the cohorts. This study was supported by National Institute of Health grants UM1 CA186107 (for NHS infrastructure), R01 CA49449 (for NHS blood collection), U01 CA176726 (for NHSII infrastructure), and R01 CA67262 (for NHSII blood collection). The content is solely the responsibility of the authors and does not necessarily represent the official views of the National Institutes of Health.

