## Supplementary information for "Trans-Ancestry GWAS of Hot Flashes Reveals Potent Treatment Target and Overlap with Psychiatric Disorders"

**Supplementary Material**

### Methods

#### Further details about data availability

Data from the UK Biobank, WHI, QIMR, NHS, and NHSII are not publicly available but may be made available upon request to qualified researchers with approved proposals. For UK Biobank, data is available upon application with information available at www.UK Biobankiobank.ac.uk. For NHS and NHS II, applications can be made to the Channing Division of Network Medicine at the Brigham and Women's Hospital and further information including the procedures to obtain and access data is described at https://www.nurseshealthstudy.org/researchers. WHI data access requests can be submitted according to established procedures (https://www.whi.org/md/working-with-whi-data), and here we used summary statistics provided by Crandall et al.

#### Additional sample characteristics

The UK Biobank is a population-based health research resource consisting of approximately 500 000 people, aged between 38 years and 73 years, who were recruited between the years 2006 and 2010 from across the UK[1]. Particularly focused on identifying determinants of human diseases in middle-aged and older individuals, participants provided a range of information (including demographics, health status, lifestyle measures, cognitive testing, personality self-report, and physical and mental health measures) via questionnaires and interviews at baseline in 2006-2010 and over time. A full description of the study design, participants and quality control methods have been described in detail previously[2] [ref]. UK Biobank received ethics approval from the Research Ethics Committee (REC reference for UK Biobank is 11/NW/0382). Our UK Biobank sample was further restricted to those with available data for Townsend Deprivation Index and, in European ancestry women, those with information on the years of birth and age at menopause.

The NHS was established in the US in 1976 when 121,700 women registered as nurses aged 30–55 years completed a mailed baseline questionnaire. The NHSII is another ongoing cohort study of US women, which enrolled 116,429 nurses aged 24-42 years in 1989. An extensive range of sociodemographic, medical, and behavioral variables have been measured in these cohorts, primarily via similar biennial questionnaires since each study's baseline. The NHS and NHSII study protocols were approved by the institutional review boards (IRBs) of the Brigham and Women's Hospital and the Harvard T.H. Chan School of Public Health.

Study participants from QIMR Berghofer were drawn from two sources (subject to genotyping and answering the relevant items): (1) Adult female immediate family of twins, who returned a self-report Health and Lifestyle questionnaire between 1989 and 1992 (total cohort size ~6,000; the twins were born up to c. 1972 and registered with the ATR) and (2) Adult female twins born up to 1943 and registered with the Australian Twin Registry (ATR), who completed a brief telephone survey in 1996 (146 twins; Wave 6 in Do et al 1998).

Our analytic samples included post-menopausal women. Menopause was assessed in 2006-2010 in UK Biobank (i.e., baseline), 1989-1992 in QIMR, 2004 in NHS, and 2017 in NHSII.[3] For UK Biobank and QIMR, we included women who underwent natural menopause (not those who reported a hysterectomy in QIMR; not those who reported a hysterectomy or bilateral oophorectomy in UK Biobank). In NHS and NHSII we included women who reported undergoing surgical menopause (gynecological surgeries: single oophorectomy, bilateral oophorectomy, oophorectomy with unknown ovaries removed, surgery with uterus removed). In NHS, 8,651 women underwent natural menopause women and 4,522 underwent surgical menopause. In NHSII, 7,129 women underwent natural menopause, and 3,947 women underwent surgical menopause.

**Additional genotyping information**
We used genetic data from five genotyping platforms for our NHS sample[4,5]: (1) Illumina HumanHap Array (n=1826 natural menopause; n=998 surgical menopause), (2) Illumina OncoArray (n=1824 natural menopause; n=840 surgical menopause), (3) Illumina HumanCore Exome Chip (n=717 natural menopause; n=475 surgical menopause), (4) Illumina OmniExpress (n=2487 natural menopause; n=1201 surgical menopause), and (5) Affymetrix 6.0 (n=1797 natural menopause; n=1008 surgical menopause). Three genotyping platforms were used in NHSII: (1) Illumina HumanHap Array (n=426 natural menopause; n=252 surgical menopause), (2) Illumina OncoArray (n=1791 natural menopause; n=869 surgical menopause), and (3) Illumina HumanCore Exome Chip (Batch 1, n=1706 natural menopausal, n=1668 surgical menopause; Batch 2, n=3206 natural menopause; n=1158 surgical menopause). Data for NHS and NHSII were: imputed to the Haplotype Reference Consortium (HRC r1.1 2016), converted to PLINK format using a dosage certainty of 0.8; restricted to participants with data for all chromosomes per genotyping platform array; restricted to SNPs not missing in more than 2% of individuals for each genotyping platform array; restricted to SNPs with minor allele frequency (MAF) above 1%; and also: deviation from the expected inbreeding coefficient (fhet $< -0.2$ or $>0.2$); Hardy Weinberg Equilibrium p-value $=1x10^{-10}$; SNPs with an info threshold of >/=0.6.

UK Biobank data was genotyped using the UK BiLEVE array and the UK Biobank axiom array. QIMR genotyping was drawn from several batches genotyped on either HapMap-based (370K, 610K, 660K) or 1000-Genomes-based (Core+Exome, PsychArray) Illumina SNP arrays. Batches were independently quality controlled by standard procedures and combined post-quality control. Imputation was run separately for the two SNP array families, for the markers passing QC in all relevant batches. A merged imputed dataset was used with a quality control filter applied to both imputation runs, and a covariate marking imputation run of origin was used in the GWAS.

### Covariates used in each GWAS

To control for potential confounding by population stratification in GWAS analyses, we included principal components of genetic variation within ancestry groups. In NHS and NHSII, the top 10 principal components were derived within European ancestry individuals previously, and we included the top 10 principal components in GWAS models. In QIMR, covariates were the top 4 principal components (in previously defined European ancestry QIMR samples) and two variables identifying the imputation run. In the UK Biobank, global principal components were calculated and plotted to determine ancestry assignments (see **Figure S1**). Cutoffs were determined for African ancestry (PC1 > .00625, PC2 > .0025, -.00125 < PC3 < .0025), East Asian ancestry (PC2 < -.01), and South Asian ancestry (.0017 < PC1 < .00425, -.00875 < PC2 < -.00375, PC3 < -.00375). For European ancestry assignments, we used the 'Genetic ethnic grouping' variable specified by the UK Biobank with individuals who self-report as 'White British' and who have very similar ancestral backgrounds according to the principal component analysis[6]. Within ancestry principal components were then calculated with the bigsnpr package in R version 4.0.2 using a subset of SNPs and all non-related participants within each ancestry and 20 PCs were included in each UK Biobank GWAS. GWAS analyses in UK Biobank was also adjusted for Townsend deprivation index and, for European ancestries, the number of years between each participant's age at menopause and 2002.

**Supplementary Figure S1. Global principal components in the UK Biobank.**
Abbreviations: EUR=European ancestry, EAS=East Asian ancestry, AFR=African ancestry, SAS=South Asian ancestry, AMR=Hispanic/Latino ancestry, PC=principal component, UKB= UK Biobank

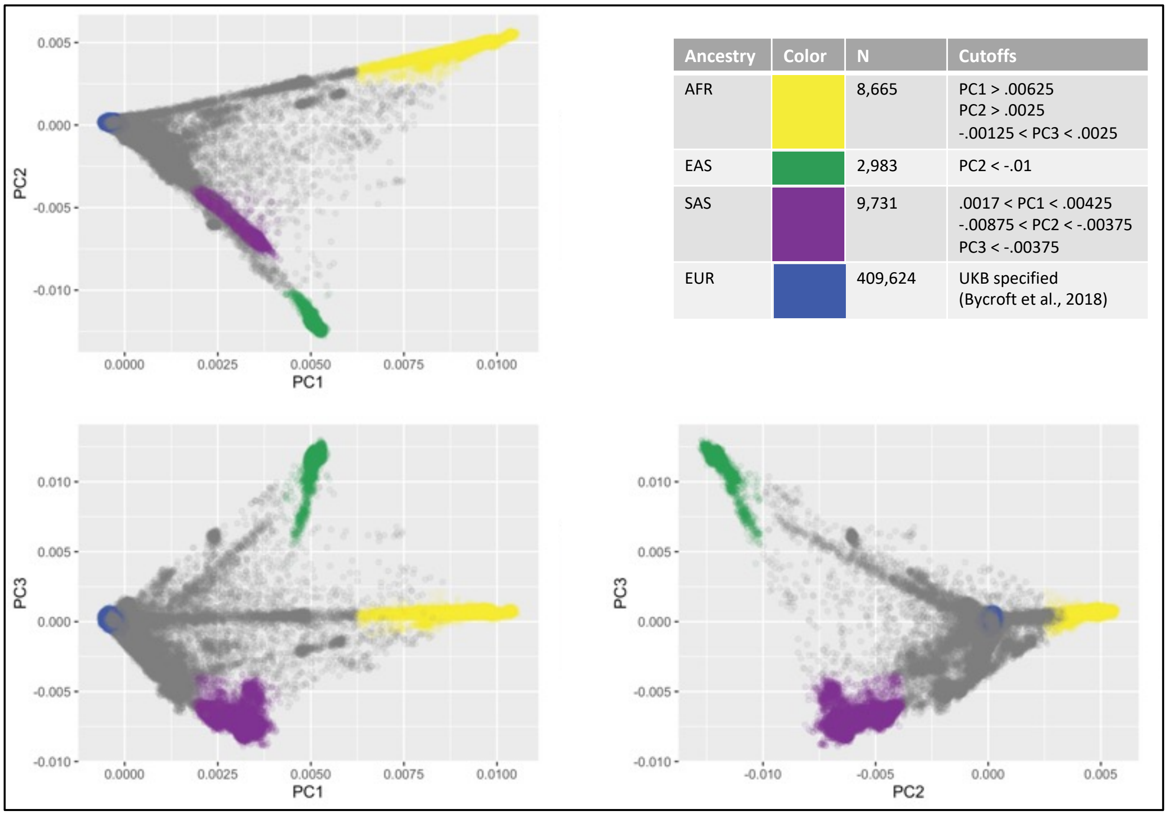


| Ancestry | Color | N | Cutoffs |
|---|---|---|---|
| AFR | | 8,665 | PC1 > .00625<br>PC2 > .0025<br>-.00125 < PC3 < .0025 |
| EAS | | 2,983 | PC2 < -.01 |
| SAS | | 9,731 | .0017 < PC1 < .00425<br>-.00875 < PC2 < -.00375<br>PC3 < -.00375 |
| EUR | | 409,624 | UKB specified<br>(Bycroft et al., 2018) |

**Statistical Analysis**

GWAS in NHS, NHSII and UK Biobank were conducted using PLINK2[7]. GWAS in QIMR was a standard 2-step analysis in SAIGE 0.45 (separately for the autosomes with LOCO, and for the X chromosome without LOCO). Step 2 was run for imputed allele dosages, restricted to minimum MAF of 0.001 and minimum MAC of 5, before post-analysis QC filtering.

Multiple rounds of meta-analyses were conducted. We ran two European ancestry meta-analyses, the first only included samples that directly measured hot flashes as the outcome phenotype (WHI, NHS, NHSII, and QIMR) whereas the second also included the UK Biobank sample in which MHT use served as a proxy for hot flashes. Results were similar regardless of whether NHS and NHSII women had undergone natural or surgical menopause, and we therefore did not distinguish between these groups for presentation.

Within African ancestry individuals, we conducted one meta-analysis that included the WHI sample from Crandall et al 2017 that directly measured hot flashes and the UKB sample that measured the proxy MHT. South Asian and East Asian samples were only available in the UKB and therefore no meta-analyses were conducted within these ancestries. We conducted three trans-ancestry meta-analyses where we included 1) all samples that directly measured hot flashes, 2) the European and African UKB samples that measured our MHT proxy, and 3) all samples of all ancestries and using either hot flashes or MHT proxy (N=149,560).

**Mendelian Randomization results overview**

None of the causal associations identified through Mendelian Randomization analyses were significant after Bonferroni correction (as stated in text, see **Supplementary Table 1** for full results). However, there were two nominally significant results for the IVW method when using genetic exposures for hot flashes, which suggested increased risk for PTSD (*beta*=.01, *se*=.003, *p*=.02) and lowered educational attainment (*beta*= -.01, *se*=.01, *p*=.01). Sensitivity analyses did not yield significant results for PTSD, indicating that the causal association between hot flashes and PTSD may not be reliable and requires further exploration. There were two nominally significant IVW results for phenotypes that putatively alter hot flash risk, with one predicted to decrease risk: Alzheimer's disease (*beta*=-.10, *se*=.05, *p*=.04), one predicted to increase risk: depression (*beta*=.25, *se*=.11, *p*=.02). Interestingly, when removing the TACR3 region from the analyses, almost all putative causal associations for the IVW method remained significant. This suggests that for most of these associations, the potential causal effect is driven from the polygenic nature of hot flashes and these disorders, rather than through the effect of NKB signaling. These results should be interpreted with caution given that we cannot be sure that all Mendelian Randomization assumptions were met.

**Supplementary Figure S2. Gene-level results for the meta-analysis of European ancestry hot flash data and Ruth et al. (2023).** The sample size for this meta-analysis was n= n=125,007. The horizontal gray line denotes Bonferroni correction for 19,171 genes tested.

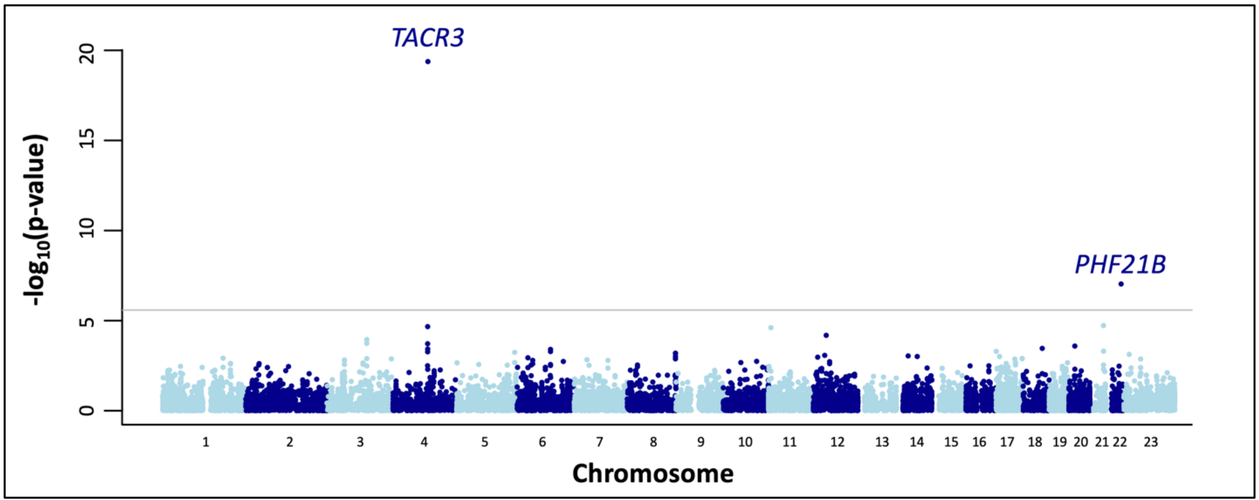
